# Evaluation of RT-PCR pooling test for the detection of SARS-CoV-2 in low-resource setting

**DOI:** 10.1101/2022.08.18.22278968

**Authors:** Narayani Maharjan, Niresh Thapa, Bibek Pun Magar, Muna Maharjan, Jiancheng Tu

## Abstract

**Background:** Specimen pooling is an efficient method when there is limited accessibility or scarcity of test kits and reagents for nucleic acid extraction and molecular detection. We evaluated the ability of the standard real-time reverse transcriptase-polymerase chain reaction (RT-PCR) test for detecting a single positive sample of severe acute respiratory syndrome coronavirus 2 (SARS-CoV-2) within a pool of negative samples and to find out the maximum pool dilution limit up to which a single positive sample can be detected.

**Methods:** RNA extracts from nasopharyngeal and oropharyngeal samples were pooled for the detection of the SARS-CoV-2 virus by RT-PCR. Positive samples were serially diluted in negative samples pools with dilutions ranging from 1/2 to 1/64 to estimate the optimal pool size. The viral transport medium (VTM) of three positive samples was also evaluated for optimal pool size determination.

**Results:** A single positive sample with a Cycle threshold (Ct) value range from 16-23 (high viral load) can be detected in dilution pools upto1/64 for both genes. In pooling before RNA extraction, a positive sample with a low Ct value (13) and intermediate Ct value (25) was detected till 1/32 dilution pool but a positive sample with a high Ct value (32) was not detected further 1/4 dilution. Besides, two positive VTM samples were detected in pools of sizes 5, 8, and 10.

**Conclusions:** This study concluded that sample testing by pooling is reliable if done properly and can help increase testing capacity in a low-resource setting.

## Background

The ongoing pandemic of the recently-emerged severe acute respiratory syndrome coronavirus 2 (SARS-CoV-2), the virus that causes coronavirus disease 2019 (COVID-19), is critically challenging health systems worldwide. Testing for SARS-CoV-2 has been limited due to the considerable strain on global supply chains for reagents, personal protective equipment, and other consumables [1,2]. The most accurate laboratory diagnosis of SARS-CoV-2 is based on nucleic acid amplification tests (NAAT) such as real-time reverse transcriptase-polymerase chain reaction (RT-PCR) [3]. Due to the rapid spread of the virus along with the increasing demand for confirmatory RT-PCR tests, the limited availability or insufficient supplies of test kits and reagents for nucleic acid extraction and molecular detection for SARS-CoV-2, has become a key bottleneck in low-and middle-income countries, as the pandemic expands [4]. The rapid diagnosis of COVID-19 in both symptomatic and asymptomatic patients can shed light on transmission patterns and facilitate contact tracing [1].

Sample pooling is a method of screening a large number of patients for infection and typically involves combining multiple patient samples into a single test sample, then testing multiple such samples [5]. Pooled nucleic acid testing has been applied in other infectious diseases like Human Immunodeficiency Virus (HIV), Hepatitis B, Hepatitis C, and influenza as a cost-effective strategy and is especially attractive as it requires no additional training, equipment, or materials. This method is first suggested by Dorfman in 1943 [6] and perfected over the years [7–9]. Samples are mixed and tested at a single pool, and subsequent individual tests are made only if the pool tests positive. It saves time, manpower, and most importantly diagnostic kits and reagents [10].

Nonetheless, as SARS-CoV-2 is a novel pathogen, it is unclear how diluting a sample containing its RNA would affect the sensitivity of this assay and the false-negative rate. This study aims to assess the ability of the standard RT-PCR test for detecting a single positive sample within a pool of negative samples and to find out the maximum pool dilution limit up to which a single positive sample can be detected.

## Methods

### Sample collection

Nasopharyngeal/Oropharyngeal swabs were collected by healthcare providers and transported in a 3ml viral transport medium (VTM) maintaining a proper cold chain and sent to the PCR laboratory of the Karnali Academy of Health Sciences (KAHS)-Teaching Hospital, Jumla. A volume of 200 microliters (μl) of the sample was further processed for nucleic acid extraction by using an automated magnetic beads method with Nucleic acid Extraction and Purification kit, according to the instruction of the ZK-96 ZhongkeBio instruments (Nanjing ZhongkeBio Medical Technology Co., Ltd, China) as per the manufacturer’s protocol in elutes of 100μl each.

### RT-PCR tests performance in the molecular laboratory

The 10μl of the extracted RNA elutes/ sample was subjected to RT-PCR for the identification and detection of SARS-CoV-2 ORF1ab gene and E gene utilizing STANDARD M nCoV Real-Time Detection kit using Real-time PCR System Accurate-96 (DLAB Scientific Co., Ltd, China). Reactions were heated to 50°C for 15 minutes for reverse transcription, denatured in 95°C for 3 minutes, 5 cycles of pre-amplification were carried out at 95°C for 5 seconds and 60°C for 40 seconds, and then 40 cycles of amplification were carried out at 95°C for 5 seconds and 60°C for 40 seconds. Fluorescence was measured using the FAM parameter for Open Reading Frame 1ab (ORF1ab) gene and HEX parameter for Envelope (E) gene as described in the interim guidelines of National Public Health Laboratories, Nepal [11]. Also, the positive and negative controls provided by the manufacturer’s kit were included in each PCR run.

### Pooling of samples prior to RT-PCR

We randomly select previously confirmed 5 positive samples and 50 negative samples. From these negative samples, 5 sets of 6 negative pools were prepared containing equal volumes of 1, 2, 4, 8,16, and 32 unique samples. One negative sample was mixed with the pool of negative samples as a control for the positive samples. Each of the 6 pools was mixed with 1 positive sample elute to make its dilution series 1/2,1/4, 1/8, 1/16, 1/32, and 1/64 dilutions. For this, 30μl of positive samples was diluted into the 1 negative sample pool of 30μl to make a 1/2 dilution, then the1/2 dilution was diluted in 2 samples negative pool samples to make a 1/4 dilution, and so on, up to 1/64 dilution for all 5 sets. After thorough mixing, 10μl of the pooled RNA was taken and 20μl of the RT-PCR reagent mix was added to each well, then RT-PCR was performed (Fig 1).

**Figure 1.**
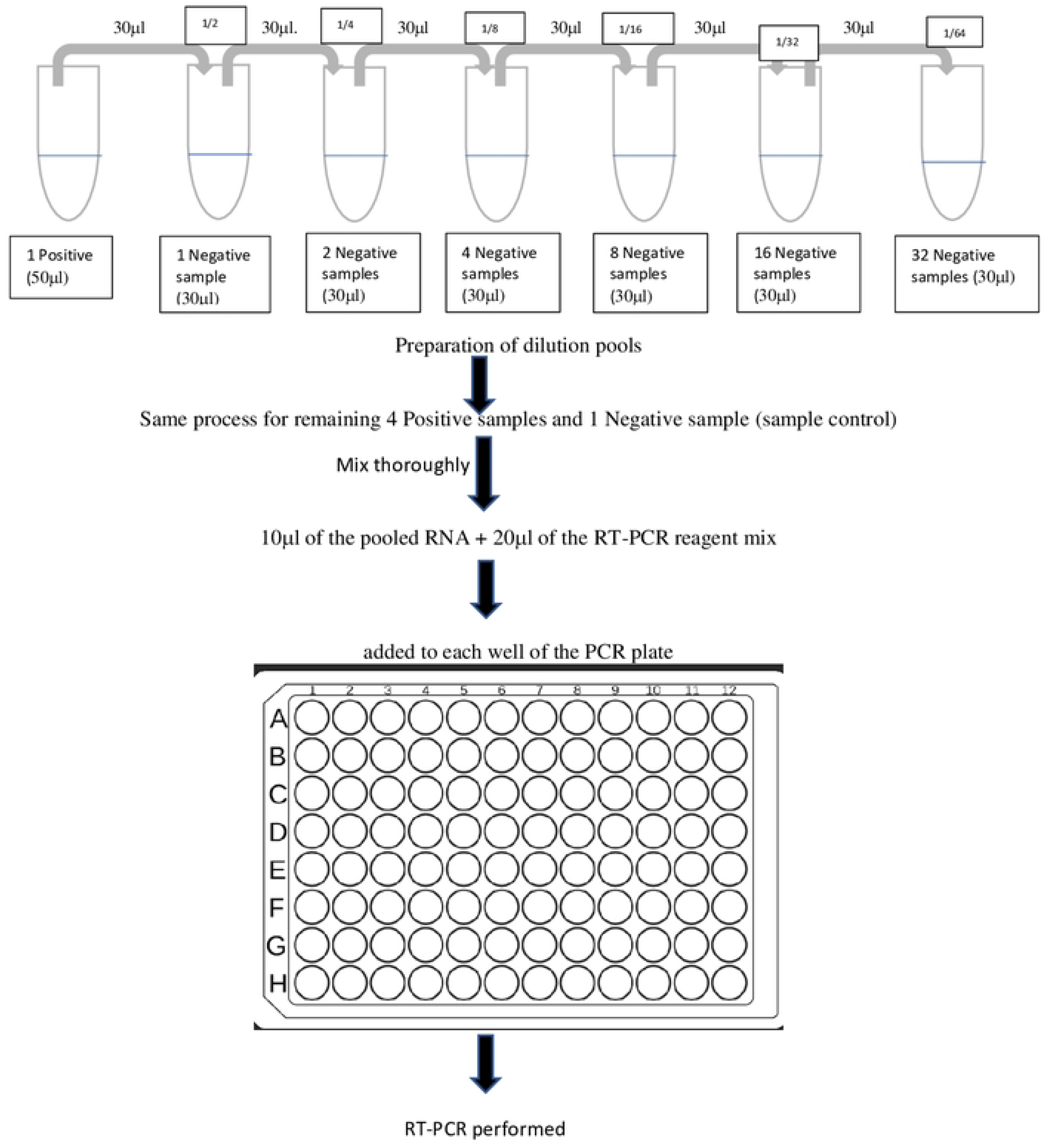
Diagrammatic representation for pooling prior to RT-PCR.

### Pooling of samples prior to RNA extraction

#### Method 1 (Pooling by serial dilution)

For this, 3 previously confirmed positive VTM samples were taken with cycle threshold (Ct) value low (16-23) and high (29-31). Then, 17 negative VTM samples were used to prepare 5 pools of 1, 2, 4, 8 and 16 negative pools with an equal volume of each negative sample. A total of 4 sets of these 5 pools of negative samples were prepared. For dilution, 200μl of the positive sample was mixed with 200μl pool of 1 negative sample to make the dilution of 1/2 and mixed thoroughly. From this 1/2 dilution, same volume that is 200μl was taken and mixed with 200μl of a pool of 2 negative samples to make the dilution of 1/4 and dilution was prepared so on up to 1/32 dilution. The remaining 17^th^ negative sample was mixed with the 4^th^ set of negative pools as a negative control for positive pools. A volume of 200μl from the single and pooled tube was mixed with of lysis buffer for inactivation, and RNA was extracted using the automated machine (ZK-96 instruments of Zhongkebio) and eluted in 100μl elution buffer. Then RT-PCR was performed for the detection of ORF1ab and E-gene.

#### Method 2 (VTM Pooling in general practice)

For this, 3 previously confirmed positive VTM samples were taken with low, middle, and high Ct values. Previously confirmed 4 negative, 7 negative, and 9 negative samples were taken to make pool size of 5, 8, and 10 samples. Then, positive samples were mixed at equal volumes with each set of negative sample pools. A volume of 200μl was pipette out from the pooled tube mixed with of lysis buffer for inactivation and RNA was extracted same as above.

### Ethical Approval

This study was approved by the Institutional Review Committee (IRC) of KAHS (IRC approval reference no 077/078/03) Jumla, Nepal.

### Statistical Analysis

Statistical analysis was performed using Microsoft Excel and the cycle thresholds (Ct) value for the amplification of each target gene were analyzed. A sample or pool was considered positive for SARS-CoV-2 if the Ct value was less than equal to 36 according to the instrument manual.

## Results

### Pooling prior to RT-PCR

In this study, previously confirmed positive samples of the SARS-CoV-2 virus were chosen to determine whether they can be detectable when the RNA elutes are mixed with negative sample elutes in different dilutions. Here a different range of Ct-value for ORF1ab and E-gene target were selected. Each series of positive samples was serially diluted with negative samples elutes up to 1/64 dilution. Then, RT-PCR was performed in the laboratory. As it was expected from the diluted sample, the amplified RNA reaches the threshold later, as the number of negative pooled samples increases. Positive samples with Ct value between the range of 16-23 (high viral load) were detected in dilution pools upto1/64 for both genes in Fig 2. But the pools containing samples with Ct value between the range of 29-30 (low viral load) were detected up to a dilution of 1/8, 1/32 for ORF1ab and ¼ and 1/16 for E-gene. The results showed that both genes were not detected in a regular pattern. But the sample with Ct value for ORF1ab/E gene 30/31 was detected till 1/64 dilution. Here for both targeted genes, the maximum Ct-value detected was between the range of 31-33. There was a gradual increase in the Ct value of an individual positive sample in the pool as the dilution increases. Thus, the probability of detection of the positive pool was decreasing as the size of the pool increased, Ct value was attained later, as we expected from dilution.

**Figure 2.**
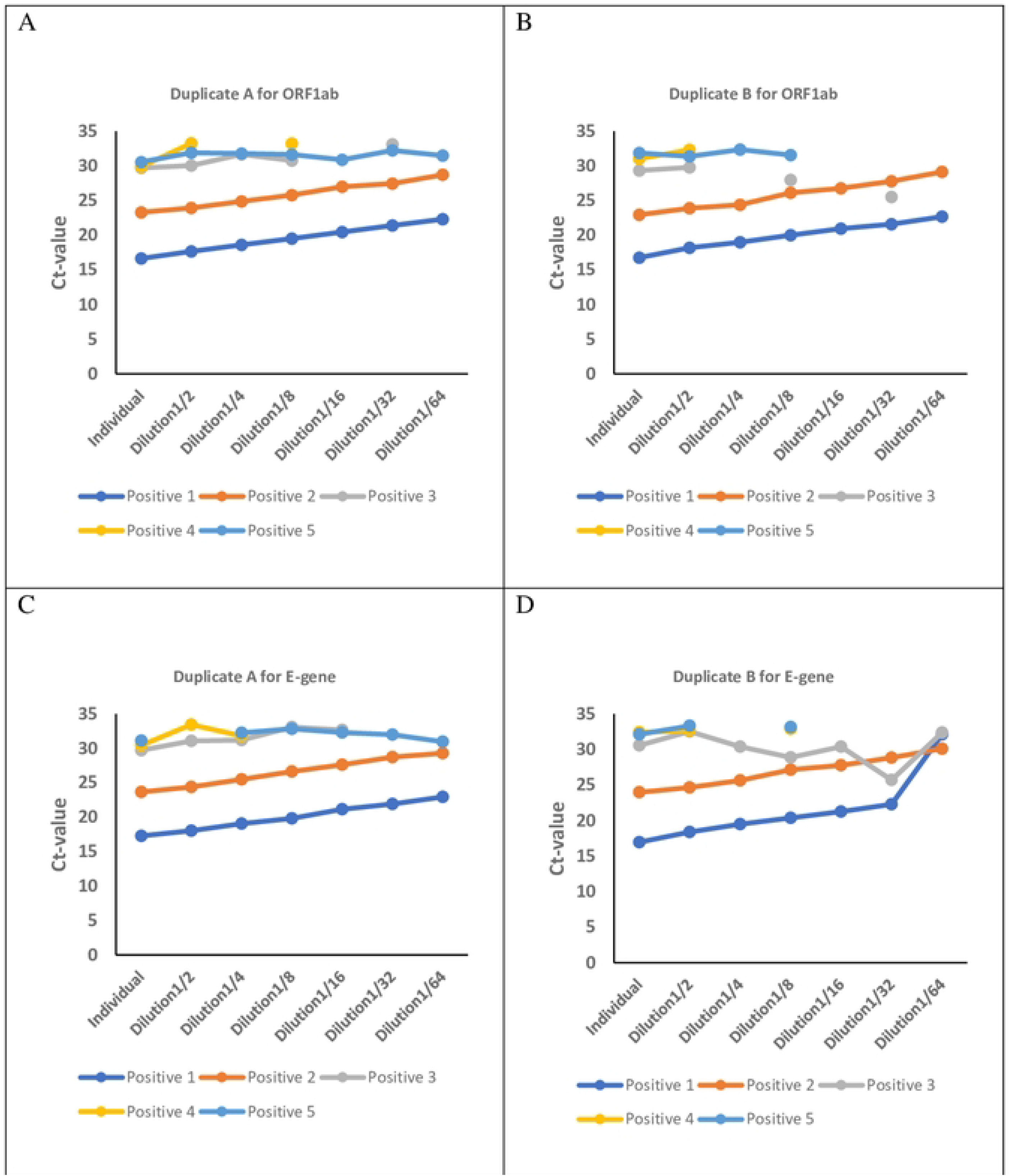
Pooling prior to RT-PCR with dilution up to 64 for each target gene. A) Duplicate A for ORF1ab; B) Duplicate B for ORF1ab; C) Duplicate A for E-gene; D) Duplicate B for E-gene. Abbreviation: Ct- Cycle threshold

### Pooling prior to RNA Extraction

#### Serial dilution of VTM samples

We performed this pooling to test whether the pooling approach can be applied even before RNA extraction, practically. Here each extracted RNA elutes of single and pooled positive samples with different Ct values were run for RT-PCR for the detection of both ORF1ab and E-gene. Positive samples with low Ct value (13) and intermediate Ct value (25) were detected till 1/32 dilution pool but the positive sample with high Ct value (32) was not detected further 1/4 dilution (Fig 3). In this pooling prior to RNA extraction, with the increase in dilution, the Ct value increases. The positive samples with high viral load can be pooled up to 32 samples or may be further.

**Figure 3.**
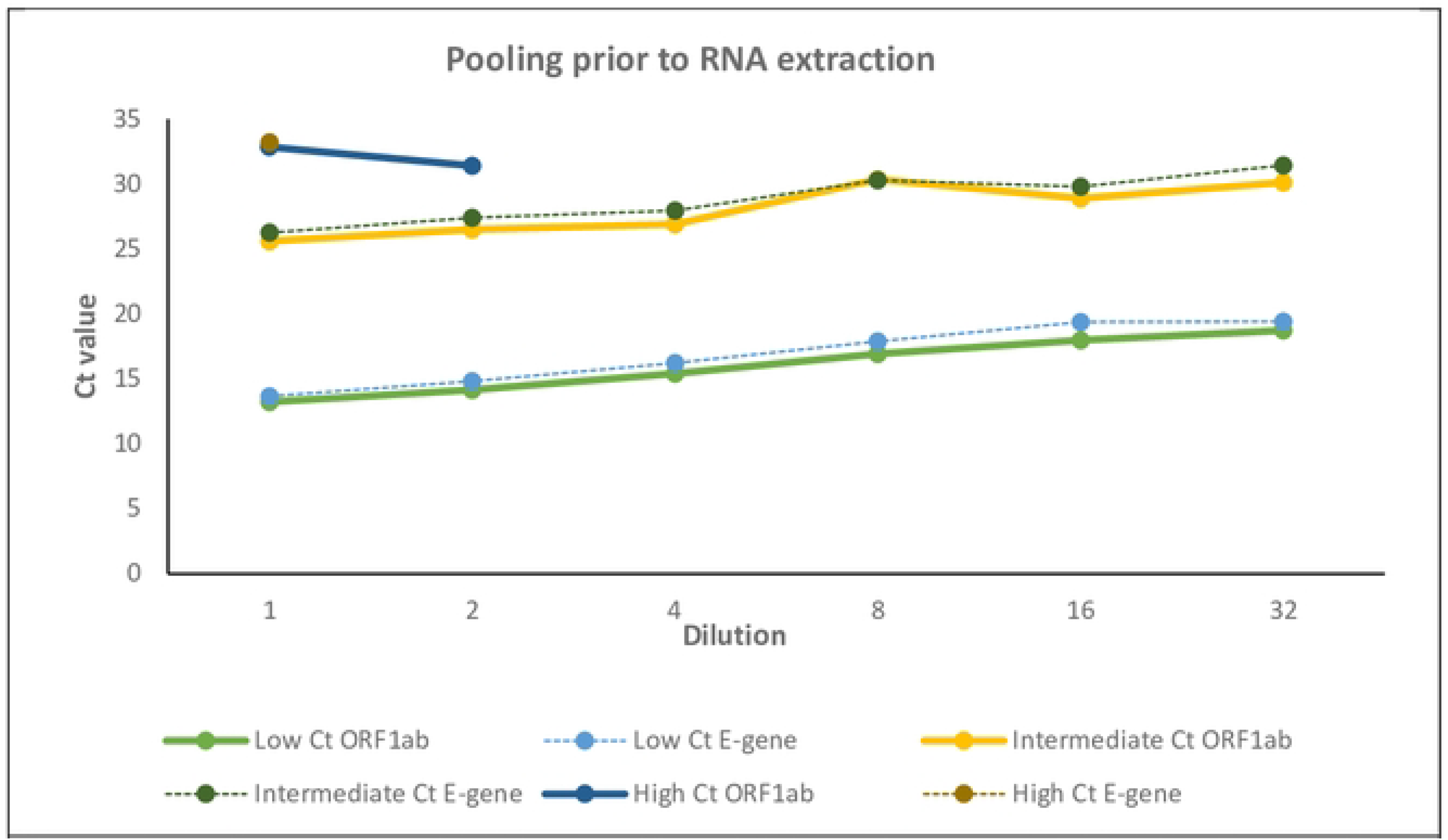
Pooling prior to RNA extraction with dilution up to 32. (Serial dilution of VTM samples). Abbreviation: Ct- Cycle threshold, VTM- Viral transport medium.

#### Direct pooling of VTM samples

The positive samples 1 and 2 were detected in all pools of size 5, 8 and 10. But for positive sample 3, ORF1ab was detected up to pool size 5. Here, the results showed that the E-gene was not detected in pool size 5 but detected in pool size 8. This showed irregularity in the detection of E-gene (Fig 4).

**Figure 4.**
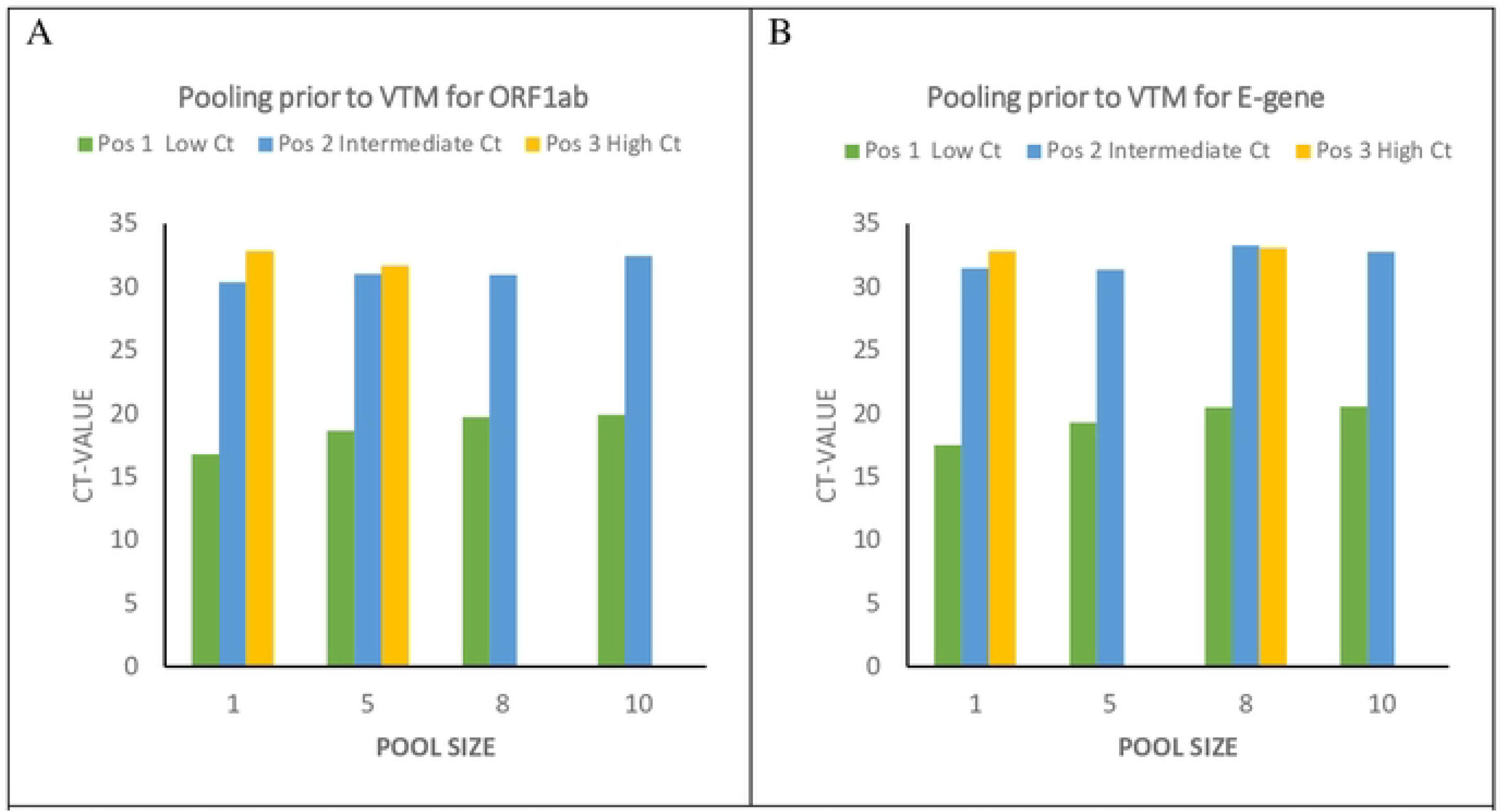
Pooling prior to VTM (Detection of positive samples with 4, 7 and 9 negative samples by direct pooling). A- Results for ORFI ab and B- Results for E-gene. Abbreviation: Ct- Cycle threshold; VTM- Viral transport 1nedium, Pos- Positive.

## Discussion

Pooling is a simple and cost-effective method, it has been used in many infectious diseases worldwide [12,13]. It saves time, work, and reagents and is found to be useful in low-resource limited countries [14]. It also allows a considerable throughput increase of clinical diagnosis and efficient for the screening of large asymptomatic populations for the presence of SARS-CoV-2 infection [15]. The present study demonstrated that pooling of SARS-CoV-2 RNA samples can be done easily in the laboratory with both pooling prior to RT-PCR and pooling prior to RNA extraction.

The study showed that the extracted RNA elutes can be detected in a pool of up to 64 samples with high viral load (Ct value 16-23) in pooling prior to RT-PCR. The result correlated with the study done in Israel where a single positive RNA elute sample could be detected in pools of up to 32 samples, with an estimated false-negative rate of 10% and showed experimentally that 64 samples could be pooled together that contains enough RNA copies for detection [4]. Also, the study results showed some irregularity/discontinuous in Ct values and the detection of targeted genes in pooled samples with high Ct-value (low viral load) between the two duplicate PCR runs which was in concordance with the results of Wacharapluesadee S. et al [5].

Further, the pooling process can be done by pooling the nasopharyngeal/ oropharyngeal swab containing VTM samples and perform RNA extraction for pooled VTM samples [16]. The major bottleneck of RNA extraction can be removed by this method [17]. Here, we also demonstrated that pooling also works well when applied before RNA extraction by serial dilution process. The positive samples with low and intermediate Ct value (high viral load) for ORF1ab and E-gene were detectable even in the dilution of 1/32. But with a high Ct value (low viral load), ORF1ab was detectable up to a dilution of ½ whereas E-gene was undetectable in any of the dilution pools. The reason may be the low sample volume or some other technical errors. This suggested that the samples with a low viral load of SARS-CoV-2 were undetectable with further dilution when pooling prior to RNA extraction.

To increase testing throughput, consuming fewer reagents and faster results yield, multiple sample pooling can be a good alternative [4,16–18]. The present gold standard for detection of SARS-CoV-2 is qPCR, which requires resources that are currently limited [5]. Hence, it is suitable for underdeveloped or developing countries, where there is a lack of resources and supplies and was proved by many studies done recently [19–24]. Thus, we performed direct pooling of VTM samples before RNA extraction as there would be very less chance in delay for detection and further decision making. It was observed that the positive sample with high viral load was again detected in all 3 negative sample pools of 5, 8 and 10. Hence, the sensitivity of detection was not affected in a pool size of 10 with a high viral load sample. This was in agreement with the studies of Chhikara K. et al and Chen F. et al [14,19]. Even a relatively weak positive sample with Ct value between 30.38-31.48 was detected in all pool size as similar to Yelin et al where they detected all three target genes (N, RdRp and E) in 8 sample pools prior to RNA extraction [4]. Similarly, a recent study also demonstrated that a single positive sample can be detected up to 7 samples pool [25]. But, in the low viral load containing positive sample pool, only one gene was detected in pool size 5 and 8 (ORF1ab and E-gene respectively) quite similar to the results obtained in pooling prior to RT-PCR. Other studies showed that 5 sample pool size had higher sensitivity compared to 10 and 15 pool size [22,26,27].

In clinical application, ORF1ab is the highest specificity confirmation target gene and considered to be less sensitive than other target genes [28]. In a study by Zhou Y. et al, the most conserved ORF1ab gene showed low sensitivity than other target genes such as N or E gene, which are less conservative but more sensitive by using different SARS-CoV-2 detection kits [29]. The present study also showed alteration in the detection of both target genes in pooled samples with low viral load. Thus, this suggested that the samples with single gene positive results should be re-examined with another highly sensitive detection kit or method to reduce the false-negative results.

Here, the difference in Ct value of individually tested RNA elutes and pooled were not that much changed as shown in Figure 2. There was a gradual increase in the threshold cycle of the pooled samples with high viral load in the original sample. Besides, the detection rate of pooled samples having higher Ct values could be increased by adding a few additional PCR cycles [4].

A limitation of this study is that we performed the test by using only one SARS-CoV-2 detection kit as available in our study setting. Besides, different VTM used during sample collection and cold chain maintenance during transportation of samples could be some other limitations of the study. Thus, for better evaluation of the pooling strategies, fresh specimens would be used, as there may be degradation of RNA in samples during the storage and thawing process. Further, we could use pooling by using different strategies and validate with different detection kits.

Even with these limitations, we found the maximum pool dilution limit up to which a single positive sample can be detected by performing different methods in a low-resource setting.

## Conclusions

Pooling of samples is a practical way for high-throughput of testing and mass screening of the population to minimize the spread of infection in the community. It is an effective method based on the prevalence rate of COVID-19 infection of specific regions. Finally, the study concluded that sample testing by pooling is reliable if done properly and perform validation of the reagents and kits for each RNA extraction and amplification in the laboratories.

## Data Availability

All data produced in the present work are contained in the manuscript.

## Acknowledgments

The authors acknowledge all the participants in the study. Also, like to thank the staffs and KAHS-Teaching Hospital Jumla, Nepal.

## Author contributions

N. M., J. C. T. and N. T. conceptualized and designed the study. N. M. and B. P. M. performed the experiments and data entry. N. M. and M. M. contributed to data analysis and interpretation. N. M. responsible for manuscript writing. N. T. and J. C. T. revised the manuscript. All authors read and approved the final manuscript.

## Disclaimer

There were no funding sources for this study.

